# Real-World Effectiveness and Safety of Avacopan in ANCA-Associated Vasculitis: A Systematic Literature Review and Meta-analysis

**DOI:** 10.64898/2026.06.17.26355805

**Authors:** Elizabeth Ibiloye, Niranjan Kathe, Kelsey Mirkovic, Coby Martin, Siyu Tu, Rachel Gamburg, Jatinder Kumar, Goutam Solanki, Zachary S. Wallace

## Abstract

**Background:** The efficacy and safety of avacopan in ANCA-associated vasculitis (AAV) has been established in randomized trials of of avacopan as a glucocorticoid (GC)-sparing therapy. However, real-world evidence (RWE) has an important role in confirming effectiveness and evaluating safety in more generalizable settings. This study aimed to synthesize RWE on the effectiveness and safety of avacopan in adults with AAV.

**Methods:** A systematic literature review and meta-analysis of non-interventional real-world studies was conducted in accordance with Preferred Reporting Items for Systematic Reviews and Meta Analyses (PRISMA) guidelines. Eligible studies included adults with AAV treated with avacopan in routine clinical practice. Pooled estimates of effectiveness and safety outcomes were calculated using random-effects meta-analyses. Primary outcomes included remission at 6 and 12 months and sustained remission at 12 months. Secondary outcomes included relapse, GC use and dosing, hepatotoxicity, infections, and treatment discontinuation. Exploratory outcomes included changes in estimated glomerular filtration rate (eGFR) and dialysis-related endpoints.

**Results:** A total of 71 studies were included and contributed to quantitative analyses. Pooled remission for patients on avacopan was 87% (95% CI: 75%-94%) at 6 months and 93% (95% CI: 86%-97%) at 12 months, and sustained remission was 86% (95% CI: 74%-93%) at 12 months. Relapse at 12 months was low (7%; 95% CI: 4%-11%). GC use was 36% at both 6 and 12 months. Improvements in eGFR were observed at 6 months (18 mL/min/1.73 m²) and 12 months (18 mL/min/1.73 m²), and dialysis liberation was 66% in a limited subset. Among avacopan patients, 11% experienced any hepatotoxicity, including 7% with serious (defined as directly reported or requiring hospitalization) hepatotoxicity, while 7% experienced serious (defined as directly reported or requiring hospitalization) infection.

**Conclusions:** In real-world clinical practice, avacopan is associated with high remission rates, low relapse rates, and a consistent GC-sparing effect, with effectiveness comparable to standard-of-care regimens. Findings support its clinical use with appropriate safety monitoring; however, the observed heterogeneity in hepatotoxicity and the limited comparative effectiveness evidence highlight areas requiring further investigation.

## Introduction

ANCA-associated vasculitis (AAV) is a rare, systemic autoimmune disease characterized by necrotizing inflammation of small- to medium-sized blood vessels, most commonly presenting as granulomatosis with polyangiitis (GPA) or microscopic polyangiitis (MPA).^1^ The disease is frequently organ- and life-threatening, with kidney involvement being common and an important contributor to morbidity and long-term outcomes.^1^ This necessitates prompt and effective remission induction to control inflammation and prevent irreversible organ damage.^2^ Standard induction regimens rely on immunosuppressive agents such as rituximab (RTX) or cyclophosphamide (CYC) in combination with high-dose glucocorticoids (GCs). Although effective for rapid disease control, prolonged GC exposure is associated with substantial toxicity, including increased risk of infection and metabolic complications.^2, 3^ Management guidelines consistently recommend minimizing GC exposure, when possible, because of the toxicity risks.^4, 5^ This creates a fundamental clinical challenge of achieving rapid and durable disease control, while minimizing the GC toxicities.

Avacopan is an oral, selective complement C5a receptor 1 (C5aR1) antagonist that inhibits C5a-mediated neutrophil activation and migration, mechanisms implicated in inflammatory processes relevant to AAV.^6^ The efficacy and safety of avacopan were established in the pivotal ADVOCATE randomized trial, in which avacopan, administered in combination with CYC (followed by azathioprine) or RTX, was noninferior to a standard prednisone taper for remission at week 26 and superior for sustained remission at week 52.^7^ In ADVOCATE, participants randomized to avacopan had an 80% reduction in median GC exposure compared to the prednisone taper group. These findings positioned avacopan as a GC-sparing therapeutic option in AAV. However, clinical trial populations are typically selected and managed under controlled conditions, whereas real-world use occurs across more heterogeneous patient populations, including those with more severe kidney involvement, and under variable monitoring practices, treatment settings, and concomitant induction strategies. Therefore, evaluations of the real-world evidence (RWE) are important to understand the effectiveness and safety of avacopan in broader populations.

The existing RWE on avacopan was previously summarized by Berke et al., which included 16 studies comprising 447 patients and primarily focused on early effectiveness and selected safety outcomes.^8^ While the synthesis included important data on outcomes such as 12-month remission and treatment discontinuation, these were reported based on limited evidence available at the time. Additional endpoints, including 12-month sustained remission, relapse, kidney outcomes, and general infections, were beyond the planned scope of that analysis. Since the analysis by Berke et al, the volume and scope of real-world data have expanded, enabling a more robust and systematic assessment of additional outcomes. The current systematic literature review and meta-analysis therefore builds on the previous study by incorporating newly published studies and conference abstracts and by evaluating a broader range of effectiveness and safety outcomes. Specifically, we evaluated remission at 6 and 12 months, sustained remission, relapse, GC use and dosing, kidney outcomes, any and serious hepatotoxicity, any and serious infections, treatment discontinuation and reasons, as well as subgroup analyses by geography (including Japan). Collectively, these additional data strengthen the assessment of both the durability of disease control and the overall benefit-risk profile of avacopan in routine clinical practice.

Building on the expanded and more comprehensive evidence base described above, available real-world data allow for a more robust assessment of both effectiveness and safety outcomes of avacopan in routine clinical practice. Prior syntheses, including Berke et al., highlighted important gaps in the available evidence, including limited evaluation of longer-term outcomes and heterogeneity in safety findings across regions, particularly with respect to hepatotoxicity as a central benefit-risk dimension, for which variable rates have been observed across real-world cohorts, including higher incidence in certain geographic regions such as Japan.^6^ Accordingly, an updated systematic literature review and meta-analysis, incorporating RWE published and presented through 28 February 2026, is warranted to synthesize non-interventional real-world evidence on the effectiveness and safety of avacopan in adults with AAV.

## Methods

### Design and reporting

This study was conducted as a systematic literature review and meta-analysis of aggregate, study-level, non-interventional RWE to evaluate the effectiveness and safety of avacopan in adults with AAV. The review was performed and reported in accordance with the Preferred Reporting Items for Systematic Reviews and Meta-Analyses (PRISMA) 2020 statement, with additional consideration of PRISMA-Harms guidance given the clinical importance of hepatotoxicity and infection outcomes.^9, 10^ Where applicable, PRISMA-S recommendations were followed to ensure transparent and reproducible reporting of the literature search strategy.^11^

This evidence synthesis was conducted under a prespecified protocol, which defined the study objectives, eligibility criteria, outcomes, and analytical approach. The protocol was prospectively registered in the International Prospective Register of Systematic Reviews (PROSPERO 2026, CRD420261365953).^12^

### Eligibility criteria

Eligibility criteria were defined using the PICOS framework (Population, Intervention, Comparator, Outcomes, Study design).^13^ Studies were eligible if they included adults (≥18 years) with GPA, MPA, or broader AAV treated with avacopan in routine clinical practice, excluding eosinophilic granulomatosis with polyangiitis (eGPA) patients. Eligible study designs included non-interventional observational studies, such as cohort studies, registries, database studies, chart reviews, and case series with ≥5 patients and extractable outcome data. Studies were considered regardless of geographic location or care setting. Conference abstracts and posters were included even in the absence of a corresponding full manuscript, provided sufficient methodological detail and outcome data were available to support data extraction and risk-of-bias assessment.

Studies were excluded if they were interventional clinical trials (except for contextual reference), case reports or case series with fewer than 5 patients, or reports without extractable data for any of the prespecified outcomes or reported in non-English languages. Given the potential for multiple publications arising from the same underlying cohort, overlapping studies were identified and managed using a prespecified hierarchy. Where linkage between reports was established, the most comprehensive and/or most recent publication was prioritized to avoid double counting, and additional reports were used to supplement missing data where appropriate.

### Information sources and search

Information sources included bibliographic databases and conference proceedings. The electronic search was conducted from October 1, 2021, through February 28, 2026, in Medical Literature Analysis and Retrieval System Online (MEDLINE), Embase and Cochrane Central Register of Controlled Trials (CENTRAL). To enhance capture of unpublished or recent evidence, conference abstracts were searched via Embase conference records and targeted searches of major rheumatology and nephrology congresses (i.e., American College of Rheumatology (ACR) Annual Meeting, American Society of Nephrology (ASN) Kidney Week, UK Kidney Week (UKKW) European Renal Association (ERA) Congress, European Alliance of Associations for Rheumatology (EULAR) Congress, European Vasculitis Society (EUVAS) meeting, International Society of Nephrology (ISN) / World Congress of Nephrology (WCN), International Vasculitis and ANCA Workshop (IVW), Japan College of Rheumatology (JCR) Annual Scientific Meeting). Reference lists of relevant reviews were hand searched, and forward citation tracking was performed where feasible. The full search strategies (including controlled vocabulary and keywords) are provided in the Supplement material.

### Study selection, extraction, and risk of bias

Title and abstract screening and full-text review were conducted by two independent reviewers, with discrepancies resolved through consensus and, if required, adjudication by a third reviewer. Data extraction was performed by one reviewer and independently verified by a second reviewer to ensure accuracy and completeness.

A standardized data extraction form was used to capture key study characteristics, including publication details, data source, geographic location, study design, baseline patient demographics, AAV subtype, ANCA status, baseline kidney involvement, avacopan treatment regimen, background immunosuppressive therapy, GC use, outcome definitions, event counts and denominators, and duration of follow-up.

Risk of bias was assessed using design-appropriate tools provided by National Heart, Lung and Blood Institute similar to Berke et al.^8, 14^ For conference abstracts and other reports with limited reporting, risk-of-bias assessment incorporated evaluation of reporting completeness and data extractability, including the availability of sufficient methodological detail and outcome information to support inclusion in quantitative synthesis. Data extraction for conference-reported evidence was based solely on information reported in the published abstract or poster.

### Outcomes

Outcomes of interest were prespecified and categorized as primary, secondary, and exploratory endpoints, based on clinical relevance and data availability. Primary effectiveness outcomes included remission at 6 months, remission at 12 months, and sustained remission at 12 months, defined as remission at both 6 and 12 months. Where multiple definitions were reported, the study-defined primary definition was used for the main analysis. Secondary outcomes included relapse at 12 months, GC use at 6 and 12 months, and cumulative GC dose (prednisone-equivalent) through 6 and 12 months. Safety outcomes included any hepatotoxicity, serious hepatotoxicity, any infection, and serious infection, based on study-reported definitions. Avacopan discontinuation was analyzed as a secondary outcome, including overall discontinuation (excluding planned stop or completion), reported reasons for discontinuation (safety and non-safety), and planned stop or completion at 6 and 12 months. Exploratory outcomes included change in eGFR from baseline at 6 and 12 months, dialysis liberation, dialysis status at 12 months, and kidney transplantation.

### Statistical analysis

For dichotomous single-arm outcomes, pooled estimates were calculated using random-effects meta-analysis of proportions, applying a logit transformation with binomial likelihood and a random study intercept to account for between-study heterogeneity.

For continuous outcomes, pooled estimates were calculated using inverse-variance random-effects models, with between-study variance estimated using the restricted maximum likelihood (REML) method.

Statistical heterogeneity was assessed using the *I²* statistic and τ*²* (tau-squared). Where heterogeneity was substantial, results were interpreted with consideration of the range and distribution of study-level estimates, rather than relying solely on pooled summary measures.

For outcomes assessed at 6 and 12 months, prespecified time windows of 4–8 months for the 6-month timepoint and 10–14 months for the 12-month timepoint were allowed.

Where necessary, summary statistics were derived from reported data (e.g., event counts from proportions), standard deviations were estimated from standard errors, confidence intervals, or other reported metrics, and medians were converted to means using established methods, in accordance with guidance from the Cochrane Handbook for Systematic Reviews of Interventions.^15^

Prespecified subgroup analyses were conducted where data permitted, including stratification by geographic region (including Japan sub-group), publication type, and induction therapy (e.g., RTX regimens with or without CYC, or other/non-specified regimens), as well as other protocol-specified study-level characteristics.

## Results

### Evidence base

A total of 781 records were identified (713 from database searches and 68 from conference proceedings). After removal of duplicates (n = 170), 611 records underwent title and abstract screening, of which 131 reports were assessed for full-text eligibility. Following full-text review, 33 reports were excluded, with reasons provided in the PRISMA flow diagram, as shown in **Figure 1**.

**Figure 1.**
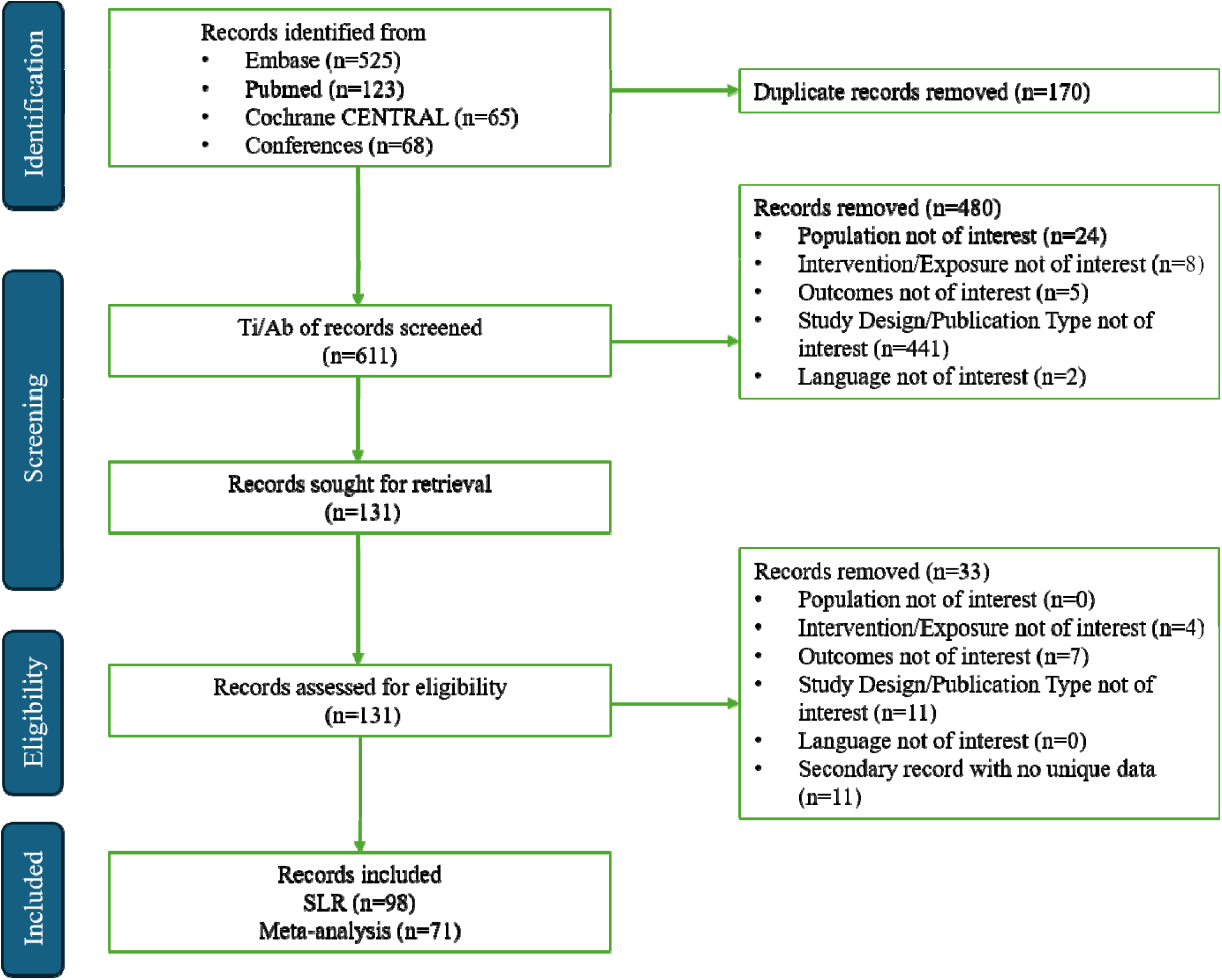
PRISMA Flow Diagram.

Overall, 71 studies were eligible for meta-analysis.^16–86^ The remaining studies were excluded from quantitative synthesis due to reasons including linkage to broader parent studies, reporting of outcomes at follow-up time points outside the prespecified time windows, and restriction to specific subgroups of the study population. Out of the 71 studies included in meta-analysis, 4 studies had good evidence quality, 53 had fair evidence quality, and 14 had poor evidence quality (Table S3).

All included studies were non-interventional RWE studies, comprising observational cohorts, registries, and case series conducted across multiple geographic regions. Studies varied in publication type, including 16 full manuscripts, 53 conference abstracts and 2 letter to the editor. A substantial proportion of studies were conducted in Japan (n = 28), which contributed notably to the safety evidence base. Full results for all outcomes evaluated in the meta-analysis are provided in Table 1. Additional study and patient characteristics are provided in the Supplementary Material.

**Table 1.** All Outcomes in Meta-Analysis.

| **Outcome** | **Result** | **95% CI** | **Studies** | **N** | **I<sup>2</sup>** | **τ<sup>2</sup>** | **p** |
| --- | --- | --- | --- | --- | --- | --- | --- |
| **Primary Outcomes** |  |  |  |  |  |  |  |
| **6-Month Remission** | **87.0%** | **75.0%, 94.0%** | **23** | **807** | **80.3%** | **2.85** | **<0.05** |
| *Regional Subgroup: Japan* | *80.0%* | *42.0%, 96.0%* | *6* | *315* | *83.7%* | *3.57* | *<0.05* |
| **12-Month Remission** | **93.0%** | **86.0%, 97.0%** | **12** | **362** | **0%** | **0.50** | **0.51** |
| *Regional Subgroup: Japan* | *90.0%* | *64.0%, 98.0%* | *3* | *157* | *0%* | *0.45* | *0.52* |
| **12-Month Sustained Remission** | **86.0%** | **74.0%, 93.0%** | **8** | **283** | **75.3%** | **0.68** | **<0.05** |
| *Regional Subgroup: Japan* | *95.0%* | *75.0%, 100.0%* | *1* | *20* | *N/A* | *N/A* | *N/A* |
| **Secondary / Exploratory Outcomes** |  |  |  |  |  |  |  |
| **12-Month Relapse** | **7.0%** | **4.0%, 11.0%** | **12** | **353** | **0%** | **0.10** | **0.60** |
| *Regional Subgroup: Japan* | *10.0%* | *4.0%, 24.0%* | *3* | *40* | *0%* | *0* | *0.82* |
| **6-Month GC Use** | **36.0%** | **21.0%, 54.0%** | **10** | **303** | **82.1%** | **1.12** | **<0.05** |
| *Regional Subgroup: Japan* | *55.0%* | *38.0%, 70.0%* | *2* | *33* | *19.4%* | *0* | *0.27* |
| **12-Month GC Use** | **36.0%** | **22.0%, 53.0%** | **7** | **191** | **79.5%** | **0.62** | **<0.05** |
| *Regional Subgroup: Japan* | *52.0%* | *30.0%, 74.0%* | *1* | *21* | *N/A* | *N/A* | *N/A* |
| **6-Month Cumulative GC Dose (prednisone-equivalent mg)** | **1743** | **1187, 2298** | **6** | **162** | **95.0%** | **405905** | **<0.05** |
| *Regional Subgroup: Japan* | *1532* | *907, 2158* | *3* | *53* | *87.0%* | *249526* | *<0.05* |
| **12-Month Cumulative GC Dose (prednisone-equivalent mg)** | **1962** | **1475, 2449** | **8** | **287** | **92.7%** | **375643** | **<0.05** |
| *Regional Subgroup: Japan* | *1940* | *1030, 2850* | *3* | *58* | *93.7%* | *580401* | *<0.05* |
| **12-Month Dialysis** | **1.0%** | **0%, 10.0%** | **2** | **68** | **0%** | **0** | **0.99** |
| *Regional Subgroup: Japan* | *N/A* | *N/A* | *N/A* | *N/A* | *N/A* | *N/A* | *N/A* |
| **6-Month Dialysis Liberation** | **66.0%** | **34.0%, 88.0%** | **4** | **28** | **0%** | **0.79** | **0.46** |
| *Regional Subgroup: Japan* | *N/A* | *N/A* | *N/A* | *N/A* | *N/A* | *N/A* | *N/A* |
| **6-Month eGFR Change Difference (mL/min/1.73 m<sup>2</sup>)** | **18.4** | **10.7, 26.2** | **2** | **35** | **3.6%** | **1.14** | **0.31** |
| *Regional Subgroup: Japan* | *N/A* | *N/A* | *N/A* | *N/A* | *N/A* | *N/A* | *N/A* |
| **12-Month eGFR Change Difference (mL/min/1.73 m<sup>2</sup>)** | **17.7** | **12.7, 22.7** | **4** | **86** | **39.0%** | **11.13** | **0.18** |
| *Regional Subgroup: Japan* | *N/A* | *N/A* | *N/A* | *N/A* | *N/A* | *N/A* | *N/A* |
| **Any Infection** | **28.0%** | **18.0%, 43.0%** | **12** | **655** | **88.0%** | **0.94** | **<0.05** |
| *Regional Subgroup: Japan* | *36.0%* | *16.0%, 62.0%* | *1* | *14* | *N/A* | *N/A* | *N/A* |
| **Serious Infection** | **7.0%** | **6.0%, 10.0%** | **28** | **1986** | **49.9%** | **0.27** | **<0.05** |
| *Regional Subgroup: Japan* | *5.0%* | *4.0%, 7.0%* | *8* | *911* | *0%* | *0* | *0.82* |
| **Any Hepatotoxicity** | **11.0%** | **7.0%, 16.0%** | **32** | **2206** | **77.9%** | **1.08** | **<0.05** |
| *Regional Subgroup: Japan* | 21.0% | 18.0%, 23.0% | 15 | 1012 | 34.5% | 0 | 0.09 |
| **Serious Hepatotoxicity** | **7.0%** | **3.0%, 18.0%** | **10** | **1239** | **85.2%** | **2.11** | **<0.05** |
| *Regional Subgroup: Japan* | 11.0% | 5.0%, 21.0% | 5 | 838 | 80.4% | 0.48 | <0.05 |
| **Avacopan Discontinuation Rates by Reason** |  |  |  |  |  |  |  |
| **Overall**<br>**[without planned stop or completion]** | **24.0%** | **20.0%, 30.0%** | **39** | **1547** | **69.6%** | **0.51** | **<0.05** |
| *Regional Subgroup: Japan* | 29.0% | 17.0%, 44.0% | 12 | 230 | 32.6% | 0.99 | 0.13 |
| **Administrative or Access** | **5.0%** | **2.0%, 11.0%** | **5** | **112** | **0%** | **0** | **0.52** |
| *Regional Subgroup: Japan* | 7.0% | 0.0%, 34.0% | 1 | 14 | N/A | N/A | N/A |
| **Hepatic Adverse Event** | **11.0%** | **6.0%, 18.0%** | **21** | **887** | **63.2%** | **1.39** | **<0.05** |
| *Regional Subgroup: Japan* | 24.0% | 12.0%, 41.0% | 9 | 145 | 0% | 1.07 | 0.76 |
| **Infection Adverse Event** | **6.0%** | **3.0%, 10.0%** | **7** | **510** | **54.7%** | **0.31** | **<0.05** |
| *Regional Subgroup: Japan* | N/A | N/A | N/A | N/A | N/A | N/A | N/A |
| **Other Adverse Event** | **9.0%** | **6.0%, 12.0%** | **22** | **1082** | **47.8%** | **0.31** | **<0.05** |
| *Regional Subgroup: Japan* | 15.0% | 10.0%, 21.0% | 6 | 157 | 0% | 0 | 0.75 |
| **Lack of Efficiency or Relapse** | **7.0%** | **4.0%, 11.0%** | **6** | **190** | **0%** | **0** | **0.91** |
| *Regional Subgroup: Japan* | 7.0% | 0.0%, 34.0% | 1 | 14 | N/A | N/A | N/A |
| **Other or Unclear** | **13.0%** | **9.0%, 20.0%** | **20** | **783** | **76.5%** | **0.82** | **<0.05** |
| *Regional Subgroup: Japan* | 8.0% | 3.0%, 17.0% | 4 | 65 | 0% | 0 | 0.84 |
| **6-Month Planned Stop or Completion** | **8.0%** | **2.0%, 28.0%** | **2** | **24** | **0%** | **0** | **0.71** |
| *Regional Subgroup: Japan* | N/A | N/A | N/A | N/A | N/A | N/A | N/A |
| **12-Month Planned Stop or Completion** | **23.0%** | **10.0%, 44.0%** | **2** | **137** | **90.3%** | **0.41** | **<0.05** |
| *Regional Subgroup: Japan* | 24.0% | 20.0%, 30.0% | N/A | N/A | N/A | N/A | N/A |

### Primary effectiveness outcomes

Pooled remission at 6 months was 87% (95% CI: 75%-94%; 23 studies; *N* = 807), with substantial heterogeneity (I² = 80.3%). In contrast, remission at 12 months was 93% (95% CI: 86%-97%; 12 studies; *N* = 362) with no observed heterogeneity (I² = 0.0%).Sustained remission at 12 months was 86% (95% CI: 74%-93%; 8 studies; *N* = 283), although estimates were heterogeneous (I² = 75.3%). Detailed estimates are provided in Figure 2.

**Figure 2.**
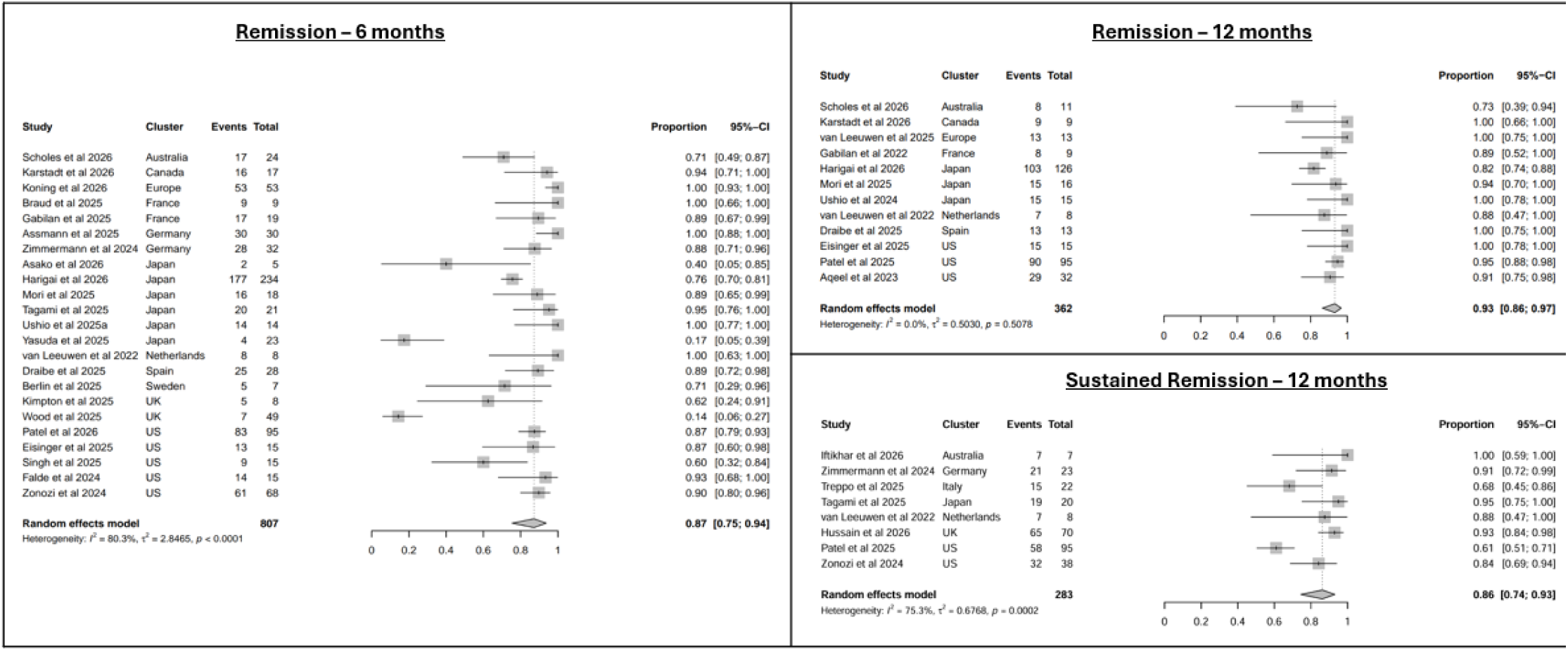
Primary Outcomes (All Regions)

### Relapse and glucocorticoid outcomes

Pooled relapse at 12 months was 7% (95% CI: 4%-11%; 12 studies; *N* = 353, I² =0.0%), indicating consistently low relapse rates across studies. Pooled estimates for GC were 36% at 6 months (95% CI: 21%-54%; 10 studies; *N* = 303, I² = 82.1%) and 36% at 12 months (95% CI: 22%-53%; 7 studies; *N* = 191, I² = 79.5%), although estimates were heterogeneous across studies.

### Kidney outcomes

In exploratory analyses, mean improvement in eGFR was 18 mL/min/1.73 m² at 6 months (95% CI: 11-26; 2 studies; *N* = 35; I² =3.6%) and 18 mL/min/1.73 m² at 12 months (95% CI: 13-23; 4 studies; *N* = 86; I² =39.0%). Dialysis liberation at 6 months was 66%, based on 28 patients across 4 studies (95% CI: 34%-88%; I² =0.0%). Only 1% of the patients were on dialysis at 12 months (95% CI: 0%-10%; 2 studies; N=68; I² =0.0%). No studies were included in the meta-analysis to allow for evaluation of kidney transplantation for kidney transplantation.

These findings should be interpreted cautiously, as kidney outcomes were derived from a limited number of studies with small sample sizes, and may be influenced by selection effects and heterogeneity in baseline disease severity and treatment practices.

### Safety and discontinuation

Pooled estimates indicated 11% (95% CI: 7%-16%; 32 studies; *N* = 2,206) for any hepatotoxicity (I² =77.9%) and 7% (95% CI: 3%-18%; 10 studies; *N* = 1,239) for serious (defined as directly reported or requiring hospitalization) hepatotoxicity (I² =85.2%), with high heterogeneity observed across studies. Any infection occurred in 28% of patients (95% CI: 18%-43%; 12 studies; *N* = 655) with high heterogeneity observed across studies (I² =88.0%). Serious (defined as directly reported or requiring hospitalization) infection occurred in 7% of patients (95% CI: 6%-10%; 28 studies; *N* = 1,986) with moderate heterogeneity (I² =49.9%).

Overall avacopan discontinuation, excluding planned treatment completion, was 24% (95% CI: 20%-30%; 39 studies; *N* = 1,547). Pooled rates for discontinuation due to specific causes were estimated independently, with hepatic adverse events at 11%, infection-related events at 6%, and other adverse events at 9%.

These findings indicate generally low to moderate rates of adverse events, although hepatotoxicity estimates were heterogeneous across studies. Importantly, pooled hepatotoxicity estimates should not be interpreted as indicating absence of risk and must be considered in the context of clinical monitoring practices and emerging postmarketing safety evidence.

### Japan subgroup

Japan contributed a substantial proportion of the included studies (28 out of 71 studies in quantitative analysis), particularly for safety outcomes (8 out of 28 studies for serious infections, 15 out of 32 studies for any hepatotoxicity, 5 out of 10 studies for serious hepatotoxicity, and 12 out of 39 studies for overall avacopan discontinuation) and was therefore examined as a prespecified subgroup. In the Japan subgroup (Figure 3, Table 1), remission at 12 months was 90% (95% CI: 64%-98%; 3 studies; *N* = 157), with a relapse rate of 10% (95% CI: 4%-24%; 3 studies; *N* = 40) and a serious infection rate of 5% (95% CI: 4%-7%; 8 studies; *N* = 911). Hepatotoxicity estimates were 21% for any hepatotoxicity (95% CI: 18%-23%; 15 studies; *N* = 1,012) and 11% for serious hepatotoxicity (95% CI: 5%-21%; 5 studies; *N* = 838). Additionally, GC use at 6 months in Japan was significantly more prevalent than that observed across all regions (55% vs 36%) (Figure 4).

**Figure 3.**
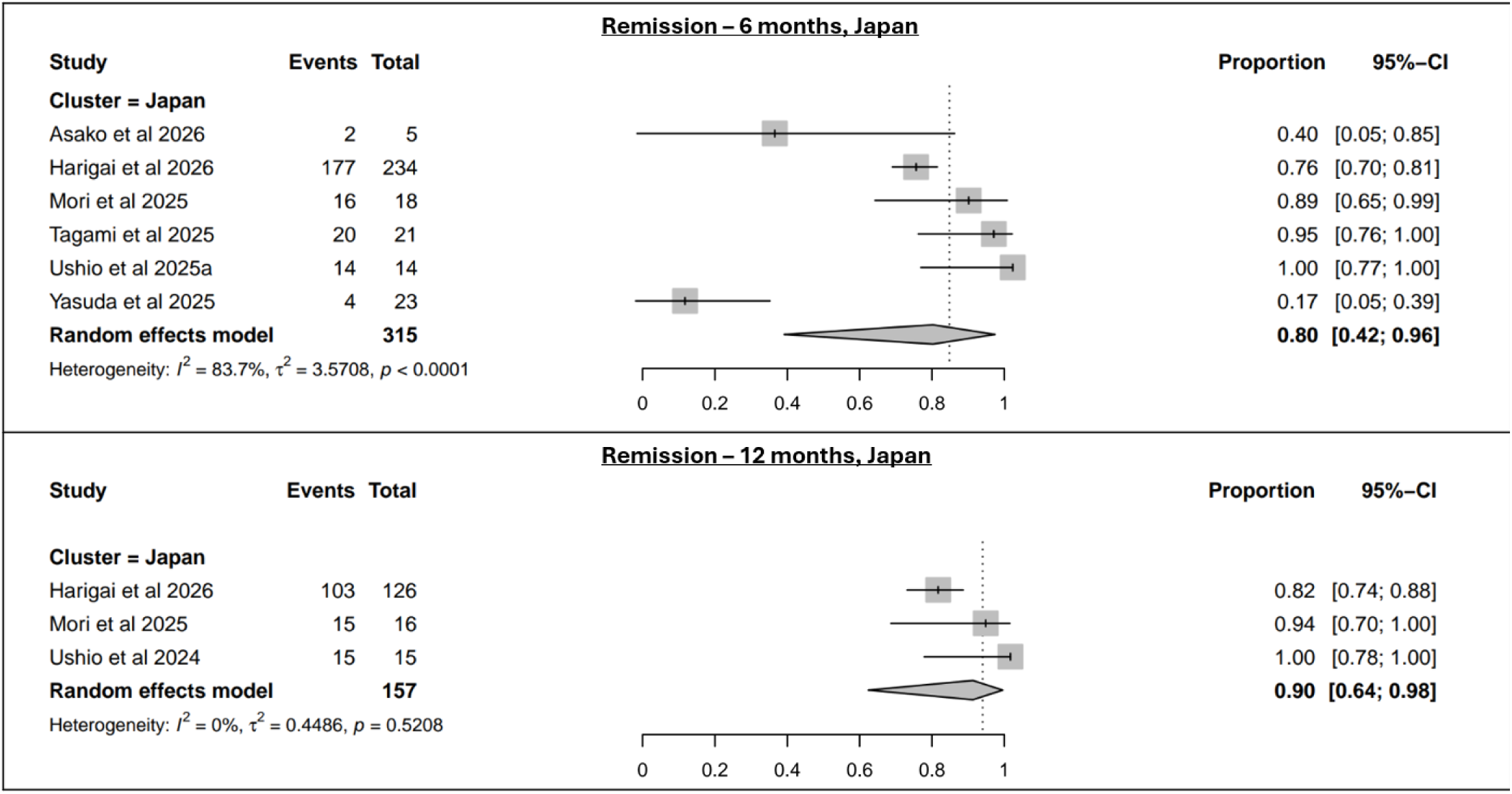
Primary Outcomes (Japan-only)

**Figure 4.**
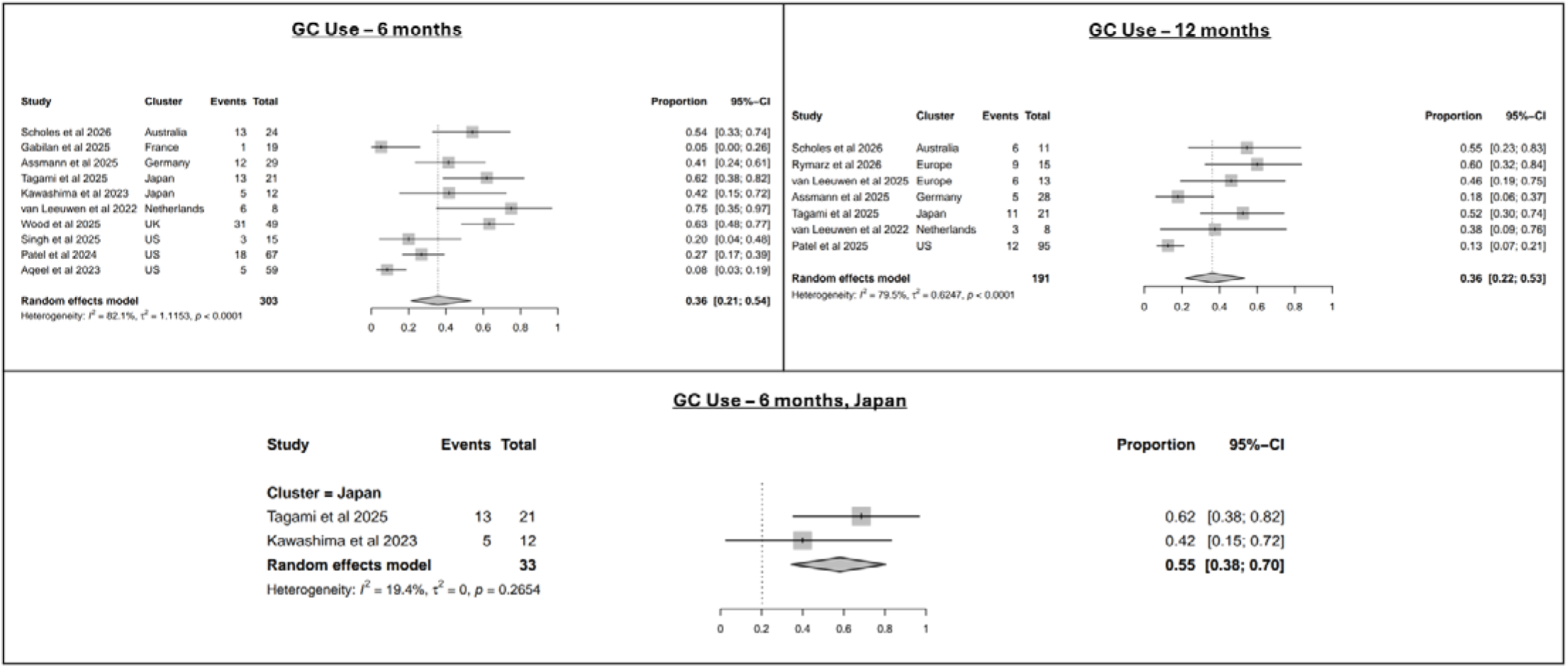
GC Use (All Regions & Japan-only)

**Figure 5.**
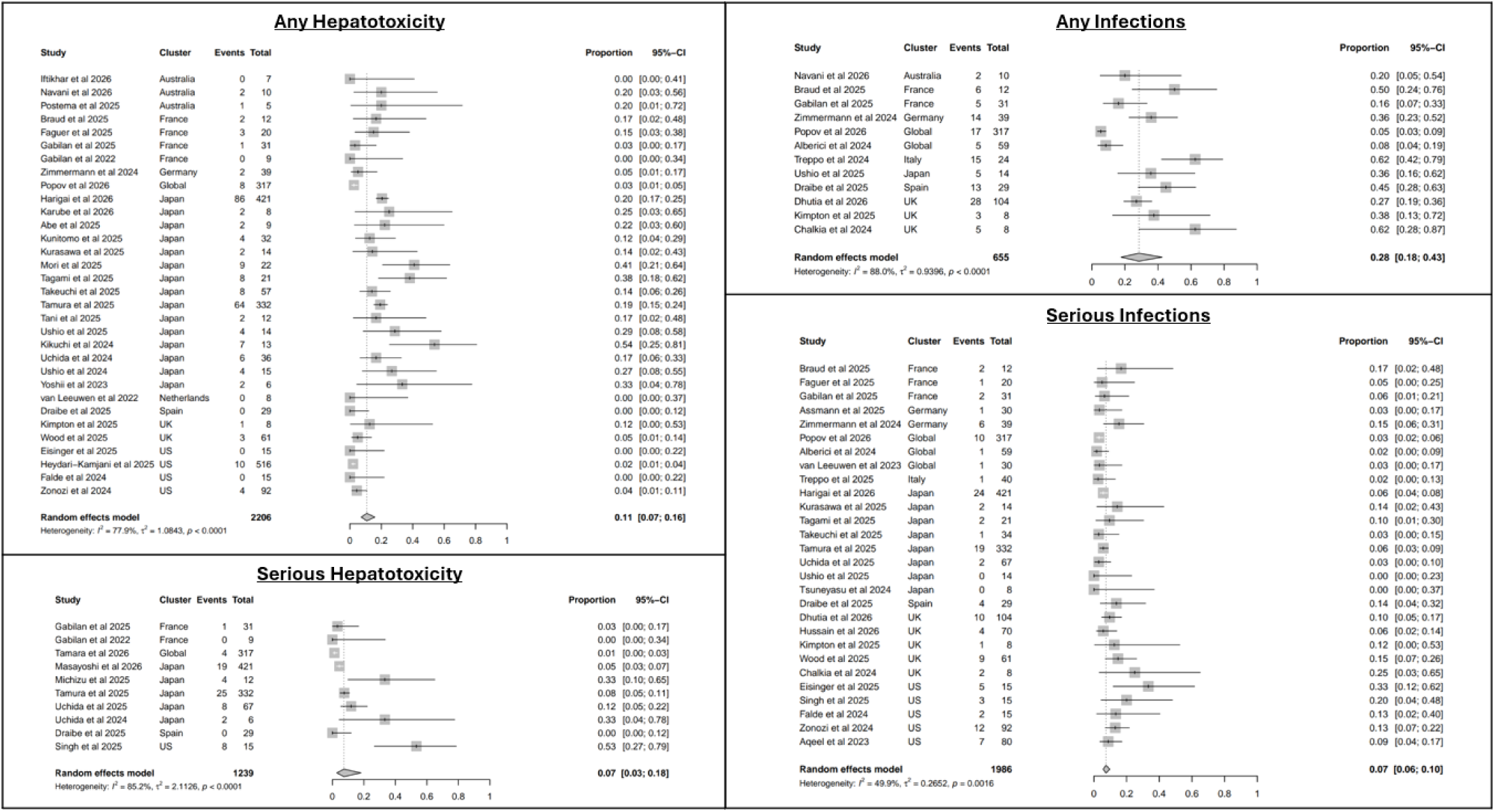
Hepatotoxicity and Infections (All Regions)

**Figure 6.**
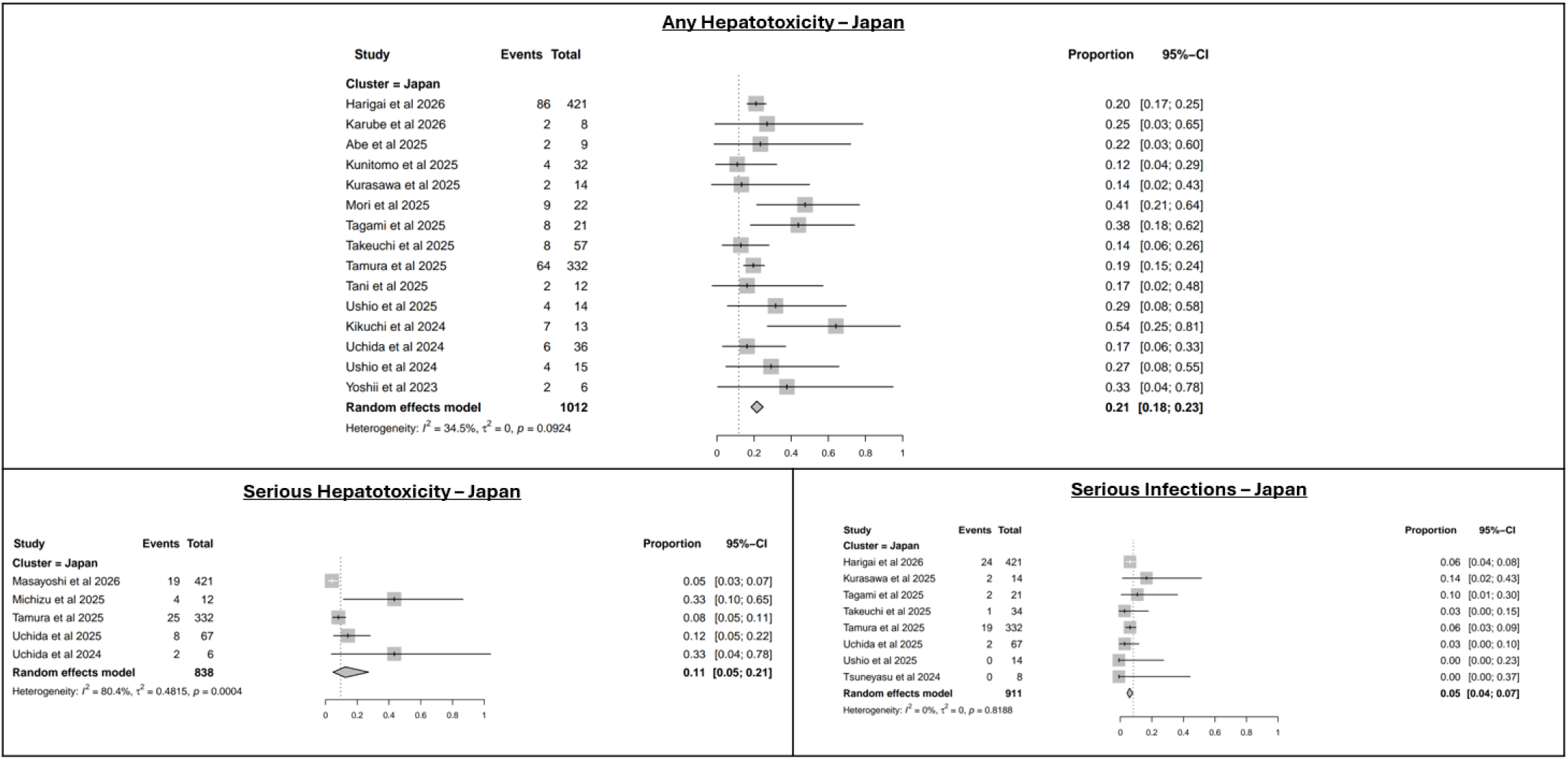
Hepatotoxicity and Infections (Japan-only)

These findings highlight regional variation in safety outcomes, particularly for hepatotoxicity, while effectiveness outcomes in Japanese cohorts were broadly consistent with overall estimates.

The results of analyses based on induction therapy subgroups can be found within the supplementary materials. Although these findings indicate generally high remission rates across induction strategies, differences should be interpreted cautiously given the observational nature of the data and potential differences in patient populations and treatment selection.

## Discussion

### Principal findings

In this systematic review and meta analysis of RWE, avacopan was associated with high remission rates at both 6 and 12 months, high sustained remission rates, and low relapse rates, indicating a favorable pattern of disease control in routine clinical practice. Remission outcomes at 12 months were consistently reported across studies, with low heterogeneity, whereas greater heterogeneity was observed at earlier timepoints and for sustained remission, likely reflecting differences in study populations, treatment approaches, and outcome definitions inherent to RWE. These effectiveness outcomes of patients treated with avacopan were accompanied by lower cumulative GC doses of 1742 mg over 6 months and 1962 mg over 12 months, compared to 2747 mg for patients receiving non-avacopan AAV treatment after 2020 and 4175 mg for patients who received treatment before 2020.^87^ Exploratory kidney outcomes suggested consistent improvement in kidney function over time and high rates of dialysis liberation compared to historic observations,^8^ although these findings were based on limited data and should be interpreted cautiously.

Safety outcomes demonstrated rates of hepatotoxicity and serious infection similar to those observed in clinical trials. Although hepatotoxicity estimates were heterogeneous across studies, with higher rates observed in the Japan subgroup. Overall, these findings suggest a favorable benefit-risk profile, characterized by sustained disease control and GC-sparing, balanced by heterogeneity in hepatic safety outcomes that warrants careful monitoring.

### Comparison with ADVOCATE and guidelines

The findings from this real-world synthesis are directionally consistent with the ADVOCATE trial.^7^ In the present analysis, remission and sustained remission rates in routine clinical practice were similarly high, supporting the external validity of randomized trial findings across more heterogeneous patient populations. The present analysis also demonstrated greater improvement in eGFR at both 6 months (18 vs 6 mL/min/1.73 m²) and 12 months (18 vs 7 mL/min/1.73 m²) compared with the ADVOCATE trial.^7^

These results are also broadly aligned with the role of avacopan in clinical practice guidance and treatment paradigms for AAV.^88^ However, given the observational nature of the included studies, these findings should be interpreted as supportive of current therapeutic positioning.

### Comparison with Berke et al

This analysis extends the prior real-world synthesis by Berke et al., which included 16 studies and 447 patients.^8^ The current analysis includes a larger evidence base and evaluates a broader range of clinically relevant outcomes, including 12-month remission, 12-month sustained remission, relapse, GC use and dosing, kidney outcomes, treatment discontinuation, and subgroup evaluations by geography (including Japan) and induction therapy.

The present findings are consistent with earlier observations of heterogeneous hepatotoxicity, particularly in Japanese cohorts, while providing a more comprehensive characterization of both effectiveness and safety outcomes across diverse real-world settings.

### Clinical interpretation

Taken together, these findings suggest that avacopan is associated with sustained disease control and meaningful reduction in GC exposure in routine clinical practice, addressing a central challenge in AAV management. The magnitude and consistency of the GC-sparing effect is particularly clinically relevant given the well-established toxicity associated with prolonged GC use. Avacopan treatment was associated with improvements in markers of kidney recovery including eGFR and dialysis-related measures, although the limited number of contributing studies and potential for selection bias preclude definitive conclusions. Overall, these results support the clinical utility of avacopan as part of induction therapy.

### Safety interpretation

Safety findings were characterized by heterogeneous estimates, particularly for hepatotoxicity, with higher rates observed in certain geographic subgroups, including Japan. Variability in hepatotoxicity estimates may reflect differences in patient characteristics, monitoring practices, and reporting patterns, and is also noted in emerging postmarketing safety data.^89^ These observations underscore the importance of careful hepatic risk assessment and liver monitoring in clinical practice, as recommended by regulatory guidance and labels worldwide.

Future research should further evaluate regional, genetic, and practice- and other patient-level factors that may contribute to heterogeneity in hepatic risk.

### Strengths and limitations

Several limitations of this study should be considered. The included studies were non-interventional and based on aggregate data, and are therefore subject to confounding, selection bias, and heterogeneity in study design and patient populations. Outcome definitions varied across studies, particularly for remission and safety endpoints, and follow-up durations were not uniform. Inclusion of conference abstracts, while improving completeness, may introduce reporting limitations, and overlapping cohorts required careful handling. In addition, small sample sizes for certain subgroups and exploratory outcomes, particularly kidney endpoints, limit precision. Finally, evidence published after 28 February 2026 is considered contextual and was not included in the primary synthesis. Nevertheless, this study has multiple key strengths. A major strength of this study is the use of a protocol-driven, systematic approach, incorporating a broad and up-to-date evidence base, including conference abstracts, and applying PRISMA-aligned methods with independent screening, extraction, and risk-of-bias assessment. The analysis includes a comprehensive set of clinically relevant endpoints, along with subgroup and sensitivity analyses to explore heterogeneity.

### Implications and future research

While these data are robust, most contributing studies lack a comparator group, highlighting the need for comprehensive real-world comparative evidence, ideally using patient-level data and standardized outcome definitions, particularly for hepatotoxicity and other safety endpoints. Additionally, longer follow-up beyond 12 months is needed to better characterize the durability of response and long-term safety profile of avacopan.

Future research should prioritize regional pharmacovigilance efforts, including comparisons between Japan and non-Japan settings, to better understand drivers of hepatic risk. Additional studies are also needed to evaluate treatment selection, monitoring practices, adherence to liver function testing, and pathways leading to treatment discontinuation. Improved characterization of kidney outcomes and their determinants in real-world populations will further inform the clinical role of avacopan.

## Conclusion

This systematic review and meta-analysis of RWE suggests that avacopan is associated with durable disease control and reduced GC exposure in routine clinical practice, with remission and relapse outcomes broadly consistent with those observed in ADVOCATE. No new safety concerns were identified, although heterogeneity in hepatotoxicity estimates, particularly across geographic regions, underscores the importance of appropriate hepatic monitoring during treatment. While kidney outcomes were suggestive of benefit in analyses, evidence remains limited and inconclusive. Overall, these findings support the clinical utility of avacopan in AAV, and support the known benefit-risk profile.

## Declarations

### Ethics approval and consent to participate

No patients or patient-level data were involved in this study, and no ethics approval or consent to participate were required.

### Consent for publication

All authors consent to the publication of this manuscript.

### Availability of data and materials

The full list of included studies within this literature review are provided within the supplementary materials. These published materials represent the sole data source for this study. Please reach out to the corresponding author for any additional inquiries.

## Funding

This study was funded by Amgen Inc.

### Authors’ contributions

All authors were involved in the conceptualization and in determining methodology for this study. Authors CM, ST, RG, JK, and GS conducted the systematic literature review, including all analyses. All authors were involved in writing and editing the manuscript.

## Acknowledgements

The authors would like to acknowledge contributions from Jared Miller and Kevin Fraccalvieri for their assistance with the systematic literature review.

## Non-standard Abbreviations and Acronyms

AAV: ANCA-associated vasculitis
AVA: Avacopan
BVAS: Birmingham Vasculitis Activity Score
CENTRAL: Cochrane Central Register of Controlled Trials
CYC: Cyclophosphamide
eGPA: Eosinophilic Granulomatosis with Polyangiitis
ERA: European Renal Association
EUVAS: European Vasculitis Society
GC: Glucocorticoid
GPA: Granulomatosis with Polyangiitis
I²: I-squared (Measure of Heterogeneity)
IVW: International Vasculitis and ANCA Workshop
JCR: Japan College of Rheumatology
MPA: Microscopic Polyangiitis
MD: Mean Difference
PRISMA: Preferred Reporting Items for Systematic Reviews and Meta Analyses
PROSPERO: International Prospective Register of Systematic Reviews
REML: Restricted Maximum Likelihood
RR: Risk Ratio
RWE: Real-world Evidence
RTX: Rituximab
UKKW: UK Kidney Week

