## Supplementary Material for "Real-World Effectiveness and Safety of Avacopan in ANCA-Associated Vasculitis: A Systematic Literature Review and Meta-analysis"

### **Table S1. Characteristics of included studies**

| **Author** | **Study Design** | **Data Source** | **Country/Region** | **Publication Type** | **Comparator Therapy** | **Sample Size (Avacopan)** | **Follow-up Duration*** |
| --- | --- | --- | --- | --- | --- | --- | --- |
| Scholes et al. 2026 | Retrospective cohort | Australian centers | Australia | Conference abstract | N/A | 31 | 238 days (IQR 127-424) |
| Iftikhar et al. 2026 | Retrospective cohort | Clinical records from Fiona Stanley Hospital | Australia | Conference abstract | N/A | 7 | 13-36 months |
| Navani et al. 2026 | Retrospective cohort | Melbourne Health and Albury‑Wodonga Health | Australia | Conference abstract | N/A | 10 | N/A |
| Postema et al. 2025 | Retrospective cohort | Medical record review | Australia | Conference abstract | N/A | 5 | 3 months |
| Karstadt et al. 2026 | Retrospective cohort | Single-centre observational cohort | Canada | Conference abstract | N/A | 45 | ≥6 months; 12 months; 14.5 months (2-24) |
| Koning et al. 2026 | Retrospective cohort | Multicenter cohort | Europe | Conference abstract | N/A | 108 | >1.5 years |
| Leeuwen et al. 2025 | Retrospective cohort | Multicenter cohort | Europe | Conference abstract | N/A | 13 | >1.5 years |
| Gabilan et al. 2025 | Retrospective cohort | Multicenter cohort | France | Full publication | N/A | 31 | ~12 months |
| Gabilan et al. 2022 | Retrospective cohort | Multicenter cohort | France | Full publication | N/A | 9 | ≥12 months |
| Braud et al. 2025 | Retrospective cohort | N/A | France | Conference abstract | N/A | 12 | 249 days (±116) |
| Faguer et al. 2025 | Retrospective cohort | Data from the University Hospital of Toulouse | France | Full Publication | GC | 20 | 6; 12 months |
| Assmann et al. 2025 | Retrospective observational study | RUB University Hospital Minden JWK and University Medical Centre Schleswig-Holstein campus Luebeck | Germany | Full publication | N/A | 30 | 49 weeks (median) |
| Zimmermann et al. 2024 | Retrospective cohort | Multicenter cohort | Germany | Full Publication | N/A | 39 | 41 weeks |
| Assmann et al. 2025 | Prospective cohort | Data from Minden University Hospital (in the Department of Rheumatology and the Department of Nephrology) | Germany | Conference abstract | N/A | 34 | 18.9 months (±3.2) |
| Alberici et al. 2024 | Retrospective cohort | CSL Vifor (Switzerland) safety database | Global | Conference abstract | N/A | 59 | >52 weeks |
| Leeuwen et al. 2023 | Retrospective cohort | Global safety database | Global | Letter to Editor | N/A | 30 | N/A |
| Popov et al. 2026 | Retrospective cohort | Registry | Global | Conference abstract | N/A | 317 | N/A |
| Treppo et al. 2025 | Prospective cohort | Multicenter cohort | Italy | Conference abstract | N/A | 40 | N/A |
| Treppo et al. 2024 | Prospective cohort | N/A | Italy | Conference abstract | N/A | 24 | 26 weeks (IQR 22.5-52) |
| Uchida et al. 2025 | Retrospective cohort | J-CANVAS: Japan Collaborative Registry of ANCA-Associated Vasculitis | Japan | Conference abstract | Non‑avacopan | 67 | 1 year (max) |
| Uchida et al. 2024a | Retrospective cohort | Single center cohort | Japan | Full Publication | N/A | 11 | N/A |
| Uchida et al. 2024b | Retrospective cohort | Three different hospitals in Japan | Japan | Full Publication | N/A | 36 | N/A |
| Tamura et al. 2025 | Retrospective cohort | N/A | Japan | Conference abstract | N/A | 332 | 2 years |
| Ushio et al. 2025 | Retrospective cohort | Kagawa University Hospital | Japan | Full Publication | N/A | 14 | 6 months |
| Ushio et al. 2025 | Retrospective cohort | Kagawa University Hospital | Japan | Conference abstract | N/A | 21 | >12 months |
| Ushio et al. 2024 | Retrospective cohort | Kagawa University Hospital | Japan | Conference abstract | N/A | 15 | 12 months |
| Tagami et al. 2025 | Retrospective cohort | Single center cohort | Japan | Full Publication | N/A | 21 | 14 months (13-18) |
| Karube et al. 2026 | Retrospective cohort | Single-centre observational cohort | Japan | Conference abstract | N/A | 8 | 12 weeks |
| Asako et al. 2026 | Retrospective cohort | Medical records from Teikyo University School of Medicine | Japan | Conference abstract | N/A | 5 | 12 months |
| Tamura et al. 2026 | Case review | Multicenter cohort | Japan | Conference abstract | N/A | 8 | N/A |
| Mori et al. 2025 | Retrospective cohort | Medical records of two core hospitals for vasculitis in Miyagi prefecture | Japan | Full Publication | N/A | 22 | N/A |
| Takeuchi et al. 2025 | Retrospective cohort | EHR | Japan | Letter to Editor | N/A | 57 | N/A |
| Yoshida et al. 2025 | Retrospective cohort | Japan Research Committee of the Ministry of Health, Labour and Welfare for Intractable Vasculitis (JPVAS) | Japan | Conference abstract | N/A | 53 | 12 months |
| Kunitomo et al. 2025 | Retrospective cohort | EHR | Japan | Conference abstract | N/A | 32 | N/A |
| Takeyama et al. 2025 | Retrospective cohort | Clinical records from Saga University Hospital | Japan | Conference abstract | N/A | 16 | 198 days |
| Abe et al. 2025 | Retrospective cohort | Single-centre observational cohort | Japan | Conference abstract | N/A | 9 | N/A |
| Yasuda et al. 2025 | Retrospective cohort | electronic medical record data | Japan | Conference abstract | N/A | 23 | 26 weeks |
| Michizu et al. 2025 | Retrospective cohort | Single center cohort | Japan | Conference abstract | N/A | 12 | N/A |
| Kurasawa et al. 2025 | Case review | Medical records | Japan | Conference abstract | N/A | 14 | N/A |
| Tani et al. 2025 | Retrospective cohort | Single center cohort | Japan | Conference abstract | N/A | 12 | N/A |
| Tsuneyasu et al. 2024 | Retrospective cohort | medical records | Japan | Conference abstract | PSL / IV CyC / RTX | 19 | N/A |
| Kikuchi et al. 2024 | Retrospective cohort | Hospital in Japan | Japan | Conference abstract | N/A | 13 | N/A |
| Yoshii et al. 2023 | Retrospective cohort | N/A | Japan | Conference abstract | high‑dose prednisone | 6 | 12 weeks (max) |
| Kawashima et al. 2023 | Retrospective cohort | EHR | Japan | Conference abstract | N/A | 7 | 12; 24 weeks |
| Mori et al. 2025 | Retrospective cohort | Japanese AAV patient registry (J-CANVAS) | Japan | Conference abstract | IV CyC or RTX | 19 | ≤48 weeks |
| Uchida et al. 2025 | Retrospective cohort | Multicenter cohort | Japan | Conference abstract | Non‑avacopan | 16 | N/A |
| Harigai et al. 2026 | Retrospective cohort | N/A | Japan | Conference abstract | N/A | 421 | 12 months |
| Rymarz et al. 2026 | Retrospective cohort | EHR | N/A | Conference abstract | N/A | 17 | ≥12 months |
| Leeuwen et al. 2022 | Case review | Chart review | Netherlands | Full publication | N/A | 8 | N/A |
| Draibe et al. 2025 | Retrospective study | Ten different hospitals in Spain | Spain | Full publication | N/A | 29 | 456.8 days (181.7) |
| Brolin et al. 2025 | Prospective cohort | Single center cohort | Sweden | Conference abstract | N/A | 7 | 6 months |
| Lindholm et al. 2025 | Case review | Multicenter cohort | Sweden | Conference abstract | N/A | 16 | N/A |
| Dhutia et al. 2026 | Retrospective cohort | Single-centre observational cohort | UK | Conference abstract | N/A | 104 | 12 months |
| Wood et al. 2025 | Retrospective cohort | Multicenter cohort | UK | Conference abstract | CyC/RTX (historical cohort) | 61 | 6 months |
| Hussain et al. 2026 | Retrospective cohort | Large UK cohort | UK | Conference abstract | high‑dose GC | 70 | 12 months |
| Kimpton et al. 2025 | Retrospective cohort | University Hospitals Bristol and Weston (UHBW) data | UK | Conference abstract | N/A | 8 | 6 months |
| Chalkia et al. 2024 | Retrospective, observational, multicenter case series | Four medical centres in the UK | UK | Full publication | N/A | 8 | 6 months |
| King et al. 2024 | Retrospective cohort | records from a tertiary vasculitis referral center in the UK | UK | Conference abstract | N/A | 30 | 26 weeks |
| Eisinger et al. 2025 | Retrospective cohort | Three academic medical centers across the United States | USA | Full Publication | N/A | 15 | 96 weeks (21) |
| Falde et al. 2024 | Retrospective cohort | Mayo Clinic sites in Rochester, Minnesota; Jacksonville, Florida; and Scottsdale, Arizona | USA | Full Publication | N/A | 15 | 17 weeks |
| Zonozi et al. 2024 | Retrospective cohort | Data from 12 academic medical centers | USA | Full Publication | N/A | 92 | 52 weeks (max) |
| Aqeel et al. 2023 | Retrospective cohort | N/A | USA | Conference abstract | N/A | 80 | 8 months |
| Sattui et al. 2025 | Retrospective cohort | Medicare Fee-for-Service (FFS) and MORE2 Registry® databases | USA | Conference abstract | N/A | 167 | 12 months |
| Patel et al. 2025 | Retrospective cohort | N/A | USA | Conference abstract | N/A | 119 | 12 months |
| Patel et al. 2024 | Retrospective cohort | Chart review | USA | Conference abstract | N/A | 67 | 6 months |
| Jaros et al. 2025 | Retrospective cohort | Excellence Network in RheumatoloGY (ENRGY) | USA | Conference abstract | N/A | 45 | 14.0 months (11.0-19.0) |
| Heydari-Kamjani et al. 2025 | Retrospective cohort | TriNetX Research Network | USA | Conference abstract | Non‑avacopan | 552 | N/A |
| Zonozi et al. 2025 | Retrospective cohort | MarketScan Commercial and Medicare Supplemental claims data | USA | Conference abstract | N/A | 95 | 381 days (222.5-588.5) |
| Singh et al. 2025 | Retrospective cohort | Two academic centers | USA | Conference abstract | N/A | 15 | 278 (267) days |
| Patel et al. 2026 | Retrospective cohort | N/A | USA | Conference abstract | N/A | 95 | N/A |
| Patel et al. 2025 | Retrospective cohort | N/A | USA | Conference abstract | N/A | 58 | 12 months |
| **Abbreviations:** CyC, cyclophosphamide; ENRGY, Excellence Network in RheumatoloGY; EHR, electronic health record; FAERS, FDA Adverse Event Reporting System; FFS, Fee-for-Service; GC, glucocorticoids; IQR, interquartile range; MORE², Medical Outcomes Research for Effectiveness and Economics Registry; N/A, not applicable; PSL, prednisolone; RTX, rituximab; SOC, standard of care | | | | | | | |

### **Table S2. Patients characteristics of avacopan groups in included studies**

| **Author & year** | **Sample Size (Avacopan)** | **Age (years)** | **Male n (%)** | **Female n (%)** | **AAV Subtype  GPA n (%)** | **AAV Subtype  MPA n (%)** | **Baseline Kidney Involvement n (%)** | **Induction Therapy (Standardized)** |
| --- | --- | --- | --- | --- | --- | --- | --- | --- |
| Scholes et al. 2026 | 31 | NA | 16 (51.6%) | 15 (48.4%) | 17 (54.8%) | NA | 18 (58.1%) | RTX |
| Iftikhar et al. 2026 | 7 | Mean: 61 | 4 (57.1%) | 3 (42.9%) | NA | NA | 7 (100%) | RTX |
| Navani et al. 2026 | 10 | Median (IQR): 61 (34–67) | 6 (60%) | 4 (40) | NA | NA | NA | RTX |
| Postema et al. 2025 | 5 | NA | NA | NA | NA | NA | NA | Unclear |
| Karstadt et al. 2026 | 45 | NA | NA | NA | NA | NA | NA | Unclear |
| Koning et al. 2026 | 108 | Median (IQR): 61 (51–73) | 56 (52.9%) | 52 (48.1%) | NA | NA | NA | RTX |
| Leeuwen et al. 2025 | 13 | NA | NA | NA | 4 (57%) | 3 (43%) | 3 (43%) | RTX |
| Gabilan et al. 2025 | 31 | Median (IQR): 72 (64–76) | 21 (68%) | 10 (32%) | NA | NA | 30 (96.8%) | RTX/CYC combined |
| Gabilan et al. 2022 | 9 | Median (IQR): 75 (34–86) | NA | NA | NA | NA | 9 (100%) | RTX |
| Braud et al. 2025 | 12 | Mean (SD): 63 (16) | 58% | 42% | NA | NA | 12 (100%) | RTX |
| Faguer et al. 2025 | 20 | Mean (SD): 61 (19) | 13 (65%) | 7 (35%) | NA | NA | 20 (100%) | RTX/CYC combined |
| Assmann et al. 2025 | 30 | Median (range): 59 (20–81) | 15 (50%) | 15 (50%) | 20 (67%) | 10 (33%) | 30 (100%) | RTX/CYC combined |
| Zimmermann et al. 2024 | 39 | Median (IQR): 64 (51–72) | 21 (54%) | 18 (46%) | NA | NA | 33 (85%) | RTX/CYC combined |
| Assmann et al. 2025 | 34 | Median (range): 58 (21–90) | 17 (50%) | 17 (50%) | 26 (76.5%) | 8 (23.5%) | NA | RTX/CYC combined |
| Alberici et al. 2024 | 59 | NA | NA | NA | NA | NA | NA | Unclear |
| Leeuwen et al. 2023 | 30 | Median: 60 | 21 (70%) | 9 (30%) | NA | NA | NA | Unclear |
| Popov et al. 2026 | 317 | NA | NA | NA | NA | NA | NA | Unclear |
| Treppo et al. 2025 | 40 | Median (IQR): 61 (52–66) | 13 (32%) | 27 (68%) | 25 (62.5%) | 15 (37.5%) | 28 (70%) | Unclear |
| Treppo et al. 2024 | 24 | Median (IQR): 60 (52–63) | 10 (42%) | 14 (58%) | 17 (70.8%) | 7 (29.2%) | 18 (75%) | RTX/CYC combined |
| Uchida et al. 2025 | 67 | Median (IQR): 71 (58.5–76.0) | 37 (50.7%) | 30 (49.3%) | 26 (38.8%) | 41 (61.2%) | NA | RTX/CYC combined |
| Uchida et al. 2024a | 11 | NA | NA | NA | NA | NA | NA | Unclear |
| Uchida et al. 2024b | 36 | Median (IQR)  Liver dysfunction group: 74.5 (68.5–79.5)  Without liver dysfunction: 76.5 (65.3–82.0) | 14 (38.9%) | 22 (61.1%) | 3 (8.3%) | 6 (83.3%) | 20 (55.5%) | RTX |
| Tamura et al. 2025 | 332 | Mean (SD): 70.2 (13.9) | 148 (44.6%) | 184 (55.4%) | 68 (20.5%) | 264 (79.5%) | NA | Unclear |
| Ushio et al. 2025 | 14 | Median (IQR): 73 (68–81.5) | 7 (50%) | 7 (50%) | 3 (21.4%) | 11 (78.6%) | 6 (42.9%) | RTX |
| Ushio et al. 2025 | 21 | Median (IQR): 73 (67–83) | NA | 9 (42.9%) | 3 (14.3%) | 18 (85.7%) | 11 (52.4%) | RTX |
| Ushio et al. 2024 | 15 | Mean (SD): 72.9 (8.3) | 8 (53.3) | 7 (46.7%) | 3 (20%) | 12 (80%) | 6 (40%) | RTX |
| Tagami et al. 2025 | 21 | Median (IQR): 77 (66–81) | 11 (52.4%) | 10 (47.6%) | 3 (14.3%) | 18 (85.7%) | 15 (71.4%) | RTX |
| Karube et al. 2026 | 8 | Median: 82 | 1 (12.5%) | 7 (87.5%) | NA | NA | NA | Unclear |
| Asako et al. 2026 | 5 | NA | NA | NA | 1 (20%) | 4 (80%) | NA | RTX |
| Tamura et al. 2026 | 8 | Median (range): 75.6 (70–81) | 2 (25%) | 6 (75%) | 1 (12.5) | 7 (87.5) | NA | Unclear |
| Mori et al. 2025 | 22 | Mean (SD): 68 (11.1) | 8 (36.4%) | 14 (63.6%) | 10 (45.5%) | 12 (54.5%) | NA | Unclear |
| Takeuchi et al. 2025 | 57 | EI group: Median (IQR): 73.0 (32–90)  AI group: 74 (46–86)  MA group: 72.5 (50–88) | 17 (29.8%) | 40 (70.2%) | 16 (28.1%) | 39 (71.9%) | NA | EI group: RTX  AI group: Unclear  MA group: unclear |
| Yoshida et al. 2025 | 53 | NA | NA | NA | NA | NA | NA | NA |
| Kunitomo et al. 2025 | 32 | Mean (SD): 70 (11.7) | 17 (53%) | 15 (47%) | 12 (37.5%) | 20 (62.5%) | NA | Unclear |
| Takeyama et al. 2025 | 16 | Median: 74 | 7 (43.8) | 9 (56.2) | 4 (25%) | 13 (81.3%) | NA | RTX |
| Abe et al. 2025 | 9 | Mean (SD): 76 (9.3) | 2 (22.2%) | 7 (77.8%) | NA | NA | NA | Unclear |
| Yasuda et al. 2025 | 23 | Mean (SD): 81 (NA) | NA | NA | 1 (4.3%) | 22 (95.7%) | NA | Unclear |
| Michizu et al. 2025 | 12 | Median: 81 | 6 (50%) | 6 (50%) | NA | NA | NA | Unclear |
| Kurasawa et al. 2025 | 14 | Mean: 74.2 | NA | NA | 3 | 2 | NA | RTX |
| Tani et al. 2025 | 12 | Mean: 76.2 | 6 (50%) | 6 (50%) | 3 (25%) | 9 (75%) | 6 (50%) | Unclear |
| Tsuneyasu et al. 2024 | 19 | NA | NA | NA | NA | Induction group: 8 (100%)  Maintenance group: 11 (100%) | NA | Other |
| Kikuchi et al. 2024 | 13 | Mean: 73.5 | NA | NA | NA | NA | Complication rate of extrarenal lesions: 46.2% | Unclear |
| Yoshii et al. 2023 | 6 | NA | NA | NA | NA | NA | NA | Unclear |
| Kawashima et al. 2023 | 7 | Mean (SD): 70.0 (12.1) | NA | NA | 3 (42.9%) | 4 (57.1%) | NA | Unclear |
| Mori et al. 2025 | 19 | Mean: 75 | 13 (68.1%) | 6 (31.90%) | NA | 16 (84.2%) | 16 (85.7%) | RTX |
| Uchida et al. 2025 | 16 | Median: 72 | NA | NA | NA | NA | NA | Unclear |
| Harigai et al. 2026 | 421 | Mean (SD): 70.7 (13.4) | 189 (44.9%) | 232 (55.1%) | 88 (20.9%) | 332 (78.9%) | NA | Unclear |
| Rymarz et al. 2026^*^ | 17 | Mean (SD): 56.2 (17.2) | 9(60%) | 6(40%) | 0.533 | 0.4 | NA | Unclear |
| Leeuwen et al. 2022 | 8 | Median (range): 46 (23–67) | 5 (62.5%) | 3 (37.5%) | NA | NA | 5 (62.5%) | RTX/CYC combined |
| Draibe et al. 2025 | 29 | Median (IQR): 56 (46.5–67.5) | 12 (41.4%) | 17 (58.6%) | 18 (62%) | 10 (34.5%) | 23 (79.31%) | RTX/CYC combined |
| Brolin et al. 2025 | 7 | Range: 25–63 | 3 (42.9%) | 4 (57.1%) | NA | NA | NA | Unclear |
| Lindholm et al. 2025 | 16 | Mean (range): 51 (16–74) | 9 (56.3%) | 6 (37.5%) | 13 (81%) | 3 (19%) | 10 (62.5%) | RTX/CYC combined |
| Dhutia et al. 2026 | 104 | Median: 66 | 53% | 47% | NA | NA | NA | Unclear |
| Wood et al. 2025 | 61 | NA | NA | NA | NA | NA | NA | RTX/CYC combined |
| Hussain et al. 2026 | 70 | NA | NA | NA | NA | NA | NA | Unclear |
| Kimpton et al. 2025 | 8 | Mean (SD): 49.5 (24.1) | 6 (75%) | 2 (25%) | 8 (100%) | NA | 3 (37.5%) | Unclear |
| Chalkia et al. 2024 | 8 | Median (range): 64 (17–80) | 38% | 63% | NA | NA | 7 (87.5%) | RTX/CYC combined |
| King et al. 2024 | 30 | NA | NA | NA | NA | NA | NA | Unclear |
| Eisinger et al. 2025 | 15 | Mean (SD): 63 (39) | 7 (47%) | 8 (53%) | NA | NA | 14 (93%) | RTX/CYC combined |
| Falde et al. 2024 | 15 | Median (IQR): 66 (52–72) | 6 (50%) | 9 (60%) | 8 (53%) | 7 (47%) | 9 (60%) | RTX/CYC combined |
| Zonozi et al. 2024 | 92 | Mean (SD): 59 (17) | 33 (36%) | 59 (64%) | NA | NA | 71 (77%) | RTX/CYC combined |
| Aqeel et al. 2023 | 80 | Mean (SD): 59 (17) | 28 (35%) | 52 (65%) | NA | NA | 95% | Unclear |
| Sattui et al. 2025 | 167 | Mean (SD): 54.5 (17.8) | 71 (42.5%) | 96 (57.5%) | NA | NA | NA | Unclear |
| Patel et al. 2025 | 119 | Mean (SD): 59.4 (16.2) | 36 (30.2%) | 83 (69.8%) | 60 (50.4%) | 59 (49.6%) | NA | RTX/CYC combined |
| Patel et al. 2024^$^ | 67 | Mean (SD): 58.7 (15.9) | 24 (30%) | 56 (70.0%) | 38 (47.5%) | 42 (52.5%) | NA | RTX/CYC combined |
| Jaros et al. 2025 | 45 | Median (IQR): 56 (43–67) | 14 (31.1%) | 31 (68.9%) | 0.644 | 0.222 | 20 (44.4%) | Unclear |
| Heydari-Kamjani et al. 2025 | 552 | Mean (SD): 56.2 (18.6) | 237 (45.9%) | 279 (54.1%) | NA | NA | NA | Unclear |
| Zonozi et al. 2025 | 95 | Mean (SD): 54 (14.3) | NA | 59 (62.1%) | 66 (69.5%) | 17 (17.9%) | 34 (35.8%) | RTX |
| Singh et al. 2025 | 15 | Mean (SD): 57 (17) | 10 (67%) | 5 (33%) | 7 (47%) | 8 (53%) | 15 (100%) | RTX/CYC combined |
| Patel et al. 2026 | 95 | Mean: 55.2 | NA | NA | 20 (46.5%) | 23 (53.5%) | NA | Unclear |
| Patel et al. 2025 | 58 | Mean (SD): 59.1 (17.4) | 23 (39.7%) | 35 (60.3%) | 22 (37.9%) | 36 (62.1%) | 58 (100%) | RTX/CYC combined |
| **Abbreviations:** AAV, ANCA‑associated vasculitis; AI, add-on induction group; ANCA, anti‑neutrophil cytoplasmic antibody; CyC, cyclophosphamide; EI, escalation-induction group; GPA, granulomatosis with polyangiitis; IQR, interquartile range; MA, maintenance-add-on group; MPA, microscopic polyangiitis; NA, not available; RTX, rituximab; SD, standard deviation  *Study with avacopan ample size of 17, baseline characteristics only reported for 15 patients with ≥18 months of follow-up  $Ony 67 patients received avacopan, baseline$Ony 67 patients received avacopan, baseline characteristics reported for 80 patients who were prescribed avacopan.  *Note: Proportions may not sum to 100% due to missing data for certain patients or because some categories are not captured in this table. Some characteristics are reported as per author reported sub-groups when calculation for the whole group is not possible.* | | | | | | | | |

### **Table S3. Risk-of-bias and reporting-completeness assessment**

| **Author** | **Year** | **Study Type** | **Publication Type** | **Evidence Quality** |
| --- | --- | --- | --- | --- |
| Alberici et al | 2024 | Cohort / Cross-sectional | Conference abstract | Fair |
| Aqeel et al | 2023 | Cohort / Cross-sectional | Conference abstract | Fair |
| Assmann et al | 2025 | Cohort / Cross-sectional | Full publication | Fair |
| Assmann et al | 2024 | Cohort / Cross-sectional | Conference abstract | Fair |
| Brolin et al | 2025 | Cohort / Cross-sectional | Conference abstract | Poor |
| Draibe et al | 2025 | Cohort / Cross-sectional | Full publication | Fair |
| Eisinger et al | 2025 | Cohort / Cross-sectional | Full Publication | Fair |
| Faguer et al | 2025 | Cohort / Cross-sectional | Full Publication | Fair |
| Falde et al | 2024 | Cohort / Cross-sectional | Full Publication | Fair |
| Gabilan et al | 2025 | Cohort / Cross-sectional | Full publication | Good |
| Gabilan et al | 2022 | Cohort / Cross-sectional | Full publication | Poor |
| Jaros et al | 2025 | Cohort / Cross-sectional | Conference abstract | Fair |
| Kawashima et al | 2023 | Cohort / Cross-sectional | Conference abstract | Poor |
| Kimpton et al | 2025 | Cohort / Cross-sectional | Conference abstract | Poor |
| Lindholm et al | 2025 | Cohort / Cross-sectional | Conference abstract | Fair |
| Mori et al | 2025 | Cohort / Cross-sectional | Full Publication | Fair |
| Mori et al | 2025 | Cohort / Cross-sectional | Conference abstract | Good |
| Patel et al | 2024 | Cohort / Cross-sectional | Conference abstract | Fair |
| Tagami et al | 2025 | Cohort / Cross-sectional | Full Publication | Fair |
| Takeuchi et al | 2025 | Cohort / Cross-sectional | Letter to Editor | Poor |
| Tamura et al | 2025 | Cohort / Cross-sectional | Conference abstract | Fair |
| Treppo et al | 2025 | Cohort / Cross-sectional | Conference abstract | Fair |
| Tsuneyasu et al | 2024 | Cohort / Cross-sectional | Conference abstract | Fair |
| Uchida et al | 2025 | Cohort / Cross-sectional | Conference abstract | Fair |
| Uchida et al | 2024 | Cohort / Cross-sectional | Full Publication | Poor |
| Uchida et al | 2024 | Cohort / Cross-sectional | Full Publication | Fair |
| Ushio et al | 2024 | Cohort / Cross-sectional | Conference abstract | Fair |
| Ushio et al | 2025 | Cohort / Cross-sectional | Full Publication | Fair |
| Leeuwen et al | 2025 | Cohort / Cross-sectional | Conference abstract | Fair |
| Leeuwen et al | 2023 | Cohort / Cross-sectional | Letter to Editor | Poor |
| Wood et al | 2025 | Cohort / Cross-sectional | Conference abstract | Fair |
| Yoshii et al | 2023 | Cohort / Cross-sectional | Conference abstract | Poor |
| Zimmermann et al | 2024 | Cohort / Cross-sectional | Full Publication | Fair |
| Zonozi et al | 2024 | Cohort / Cross-sectional | Full Publication | Fair |
| Heydari-Kamjani et al | 2025 | Cohort / Cross-sectional | Conference abstract | Fair |
| Zonozi et al | 2025 | Cohort / Cross-sectional | Conference abstract | Fair |
| Patel et al | 2025 | Cohort / Cross-sectional | Conference abstract | Fair |
| Sattui et al | 2025 | Cohort / Cross-sectional | Conference abstract | Fair |
| Braud et al | 2025 | Cohort / Cross-sectional | Conference abstract | Fair |
| Patel et al | 2025 | Cohort / Cross-sectional | Conference abstract | Fair |
| Treppo et al | 2024 | Cohort / Cross-sectional | Conference abstract | Fair |
| King et al | 2024 | Cohort / Cross-sectional | Conference abstract | Poor |
| Kikuchi et al | 2024 | Cohort / Cross-sectional | Conference abstract | Fair |
| Patel et al | 2026 | Cohort / Cross-sectional | Conference abstract | Fair |
| Scholes et al | 2026 | Cohort / Cross-sectional | Conference abstract | Fair |
| Popov et al | 2026 | Cohort / Cross-sectional | Conference abstract | Fair |
| Harigai et al | 2026 | Cohort / Cross-sectional | Conference abstract | Fair |
| Hussain et al | 2026 | Cohort / Cross-sectional | Conference abstract | Fair |
| Dhutia et al | 2026 | Cohort / Cross-sectional | Conference abstract | Fair |
| Karube et al | 2026 | Cohort / Cross-sectional | Conference abstract | Poor |
| Karstadt et al | 2026 | Cohort / Cross-sectional | Conference abstract | Fair |
| Koning et al | 2026 | Cohort / Cross-sectional | Conference abstract | Good |
| Iftikhar et al | 2026 | Cohort / Cross-sectional | Conference abstract | Poor |
| Navani et al | 2026 | Cohort / Cross-sectional | Conference abstract | Poor |
| Yoshida et al | 2025 | Cohort / Cross-sectional | Conference abstract | Fair |
| Kunitomo et al | 2025 | Cohort / Cross-sectional | Conference abstract | Fair |
| Takeyama et al | 2025 | Cohort / Cross-sectional | Conference abstract | Fair |
| Abe et al | 2025 | Cohort / Cross-sectional | Conference abstract | Poor |
| Uchida et al | 2025 | Cohort / Cross-sectional | Conference abstract | Fair |
| Yasuda et al | 2025 | Cohort / Cross-sectional | Conference abstract | Fair |
| Ushio et al | 2025 | Cohort / Cross-sectional | Conference abstract | Fair |
| Michizu et al | 2025 | Cohort / Cross-sectional | Conference abstract | Poor |
| Tani et al | 2025 | Cohort / Cross-sectional | Conference abstract | Fair |
| Singh et al | 2025 | Cohort / Cross-sectional | Conference abstract | Fair |
| Chalkia et al | 2024 | Case Series | Full publication | Good |
| Postema et al | 2025 | Case Series | Conference abstract | Fair |
| Leeuwen et al | 2022 | Case Series | Full publication | Fair |
| Rymarz et al | 2026 | Case Series | Conference abstract | Fair |
| Asako et al | 2026 | Case Series | Conference abstract | Fair |
| Tamura et al | 2026 | Case Series | Conference abstract | Fair |
| Kurasawa et al | 2025 | Case Series | Conference abstract | Fair |

### **Table S4. Subgroup analysis – Adjunctive Therapy**

| **Outcome** | **Population** | **Result** | **95% CI** | **Studies** | **N** | **I²** | **τ²** | **p** |
| --- | --- | --- | --- | --- | --- | --- | --- | --- |
| **Primary Outcomes** | | | | | | | | |
| **6-Month Remission** | RTX | 0.92 | 0.57, 0.99 | 5 | 73 | 35.6% | 3.31 | 0.18 |
|  | RTX/CYC Combined | 0.87 | 0.71, 0.95 | 11 | 374 | 86.8% | 2.31 | <0.05 |
|  | Unclear/Other | 0.83 | 0.51, 0.96 | 7 | 360 | 78.5% | 3.47 | <0.05 |
| **12-Month Remission** | RTX | 0.91 | 0.75, 0.97 | 4 | 51 | 0% | 0.39 | 0.53 |
|  | RTX/CYC Combined | 0.95 | 0.90, 0.98 | 4 | 131 | 0% | 0 | 0.88 |
|  | Unclear/Other | 0.90 | 0.70, 0.97 | 4 | 180 | 0% | 0.32 | 0.70 |
| **12-Month Sustained Remission** | RTX | 0.96 | 0.78, 0.99 | 2 | 27 | 0% | 0 | 0.99 |
|  | RTX/CYC Combined | 0.81 | 0.63, 0.91 | 4 | 164 | 75.4% | 0.45 | <0.05 |
|  | Unclear/Other | 0.85 | 0.61, 0.95 | 2 | 92 | 86.9% | 0.63 | <0.05 |
| **Secondary Outcomes** | | | | | | | | |
| **12-Month Relapse** | RTX | 0.10 | 0.05, 0.20 | 6 | 69 | 0% | 0 | 0.94 |
|  | RTX/CYC Combined | 0.06 | 0.03, 0.11 | 5 | 214 | 10% | 0.13 | 0.35 |
|  | Unclear/Other | 0.06 | 0.02, 0.14 | 1 | 70 | N/A | N/A | N/A |
| **6-Month GC Use** | RTX | 0.58 | 0.43, 0.71 | 2 | 45 | 0% | 0 | 0.60 |
|  | RTX/CYC Combined | 0.35 | 0.17, 0.58 | 6 | 187 | 81.1% | 1.09 | <0.05 |
|  | Unclear/Other | 0.18 | 0.05, 0.49 | 2 | 71 | 86.6% | 0.79 | <0.05 |
| **12-Month GC Use** | RTX | 0.51 | 0.37, 0.65 | 3 | 45 | 0% | 0 | 0.91 |
|  | RTX/CYC Combined | 0.15 | 0.10, 0.22 | 3 | 131 | 40% | 0 | 0.19 |
|  | Unclear/Other | 0.60 | 0.32, 0.84 | 1 | 15 | N/A | N/A | N/A |
| **6-Month Cumulative GC Dose** | RTX | 1812 | 1057, 2567 | 4 | 66 | 87.7% | 472999.17 | <0.05 |
|  | RTX/CYC Combined | 1659 | 487, 2831 | 2 | 96 | 98.4% | 703533.14 | <0.05 |
|  | Unclear/Other | N/A | N/A | N/A | N/A | N/A | N/A | N/A |
| **12-Month Cumulative GC Dose** | RTX | 2004 | 1307, 2702 | 5 | 71 | 88.8% | 427319.43 | <0.05 |
|  | RTX/CYC Combined | 1929 | 1110, 2748 | 3 | 216 | 95.5% | 497127.07 | <0.05 |
|  | Unclear/Other | N/A | N/A | N/A | N/A | N/A | N/A | N/A |
| **12-Month Dialysis** | RTX | N/A | N/A | N/A | N/A | N/A | N/A | N/A |
|  | RTX/CYC Combined | 0.01 | 0.00, 0.10 | 2 | 68 | 0% | 0 | 0.99 |
|  | Unclear/Other | N/A | N/A | N/A | N/A | N/A | N/A | N/A |
| **6-Month Dialysis Liberation** | RTX | N/A | N/A | N/A | N/A | N/A | N/A | N/A |
|  | RTX/CYC Combined | 0.66 | 0.34, 0.88 | 4 | 28 | 0% | 0.79 | 0.46 |
|  | Unclear/Other | N/A | N/A | N/A | N/A | N/A | N/A | N/A |
| **6-Month eGFR Change Difference (mL/min/1.73 m²)** | RTX | N/A | N/A | N/A | N/A | N/A | N/A | N/A |
|  | RTX/CYC Combined | 18.45 | 10.68, 26.21 | 2 | 35 | 3.6% | 1.14 | 0.31 |
|  | Unclear/Other | N/A | N/A | N/A | N/A | N/A | N/A | N/A |
| **12-Month eGFR Change Difference (mL/min/1.73 m²)** | RTX | N/A | N/A | N/A | N/A | N/A | N/A | N/A |
|  | RTX/CYC Combined | 17.68 | 12.65, 22.70 | 4 | 86 | 39.0% | 11.13 | 0.18 |
|  | Unclear/Other | N/A | N/A | N/A | N/A | N/A | N/A | N/A |
| **Any Infection** | RTX | 0.36 | 0.22, 0.53 | 3 | 36 | 1.3% | 0 | 0.36 |
|  | RTX/CYC Combined | 0.38 | 0.26, 0.52 | 6 | 235 | 72.4% | 0.35 | <0.05 |
|  | Unclear/Other | 0.09 | 0.03, 0.24 | 3 | 384 | 79% | 0.34 | <0.05 |
| **Serious Infection** | RTX | 0.07 | 0.04, 0.15 | 5 | 95 | 0% | 0 | 0.65 |
|  | RTX/CYC Combined | 0.11 | 0.08, 0.15 | 13 | 526 | 29.2% | 0.10 | 0.15 |
|  | Unclear/Other | 0.05 | 0.04, 0.06 | 9 | 1357 | 0% | 0 | 0.44 |
| **Any Hepatotoxicity** | RTX | 0.18 | 0.13, 0.25 | 10 | 195 | 0% | 0.03 | 0.66 |
|  | RTX/CYC Combined | 0.04 | 0.02, 0.07 | 9 | 310 | 0% | 0 | 0.89 |
|  | Unclear/Other | 0.15 | 0.08, 0.27 | 13 | 1701 | 89.2% | 1.23 | <0.05 |
| **Serious Hepatotoxicity** | RTX | 0.10 | 0.01, 0.65 | 2 | 15 | 0% | 1.46 | 0.99 |
|  | RTX/CYC Combined | 0.08 | 0.01, 0.40 | 4 | 142 | 80.2% | 3.33 | <0.05 |
|  | Unclear/Other | 0.06 | 0.02, 0.17 | 4 | 1082 | 87.8% | 1.26 | <0.05 |
| **Avacopan Discontinuation Rates by Reason** | | | | | | | | |
| **Overall [without planned stop or completion]** | RTX | 0.32 | 0.24, 0.41 | 8 | 170 | 26.7% | 0.03 | 0.22 |
|  | RTX/CYC Combined | 0.24 | 0.18, 0.31 | 13 | 478 | 55.6% | 0.23 | <0.05 |
|  | Unclear/Other | 0.22 | 0.15, 0.33 | 18 | 899 | 78.7% | 0.94 | <0.05 |
| **Administrative or Access** | RTX | N/A | N/A | N/A | N/A | N/A | N/A | N/A |
|  | RTX/CYC Combined | 0.08 | 0.03, 0.18 | 3 | 53 | 0% | 0 | 0.47 |
|  | Unclear/Other | N/A | N/A | N/A | N/A | N/A | N/A | N/A |
| **Hepatic Adverse Event** | RTX | 0.19 | 0.12, 0.30 | 5 | 77 | 0% | 0 | 0.49 |
|  | RTX/CYC Combined | 0.05 | 0.03, 0.08 | 7 | 367 | 0% | 0 | 0.57 |
|  | Unclear/Other | 0.17 | 0.06, 0.41 | 9 | 443 | 73.5% | 2.83 | <0.05 |
| **Infection Adverse Event** | RTX | N/A | N/A | N/A | N/A | N/A | N/A | N/A |
|  | RTX/CYC Combined | 0.10 | 0.05, 0.19 | 3 | 81 | 16.3% | 0.02 | 0.30 |
|  | Unclear/Other | 0.04 | 0.02, 0.06 | 3 | 416 | 58.4% | 0 | 0.09 |
| **Other Adverse Event** | RTX | 0.09 | 0.04, 0.17 | 4 | 81 | 0% | 0 | 1.00 |
|  | RTX/CYC Combined | 0.12 | 0.09, 0.15 | 7 | 355 | 5.7% | 0 | 0.38 |
|  | Unclear/Other | 0.08 | 0.04, 0.13 | 11 | 646 | 67.6% | 0.58 | <0.05 |
| **Lack of Efficiency or Relapse** | RTX | N/A | N/A | N/A | N/A | N/A | N/A | N/A |
|  | RTX/CYC Combined | 0.05 | 0.02, 0.14 | 2 | 61 | 0% | 0 | 0.58 |
|  | Unclear/Other | 0.08 | 0.04, 0.14 | 3 | 115 | 0% | 0 | 0.71 |
| **Other or Unclear** | RTX | 0.25 | 0.13, 0.41 | 4 | 107 | 60.6% | 0.30 | 0.05 |
|  | RTX/CYC Combined | 0.13 | 0.06, 0.24 | 8 | 313 | 72.6% | 0.89 | <0.05 |
|  | Unclear/Other | 0.11 | 0.05, 0.20 | 8 | 363 | 81.1% | 0.73 | <0.05 |
| **6-Month Planned Stop or Completion** | RTX | 0.11 | N/A | 1 | 9 | N/A | N/A | N/A |
|  | RTX/CYC Combined | 0.07 | N/A | 1 | 15 | N/A | N/A | N/A |
|  | Unclear/Other | N/A | N/A | N/A | N/A | N/A | N/A | N/A |
| **12-Month Planned Stop or Completion** | RTX | N/A | N/A | N/A | N/A | N/A | N/A | N/A |
|  | RTX/CYC Combined | 0.13 | N/A | 1 | 92 | N/A | N/A | N/A |
|  | Unclear/Other | 0.38 | N/A | 1 | 45 | N/A | N/A | N/A |
| **Abbreviations:** CyC, cyclophosphamide; eGFR, estimated glomerular filtration rate; I², inconsistency statistic; τ², between-study variance; N/A, not applicable; RTX, rituximab | | | | | | | | |

### **Table S5. Embase search strategy**

| **Line** | **Search string** | **Hits** |
| --- | --- | --- |
| 1 | ('avacopan'/exp OR avacopan:ti,ab OR tavneos:ti,ab OR ccx168:ti,ab OR ('c5a' NEAR/3 receptor NEAR/3 antagonist*):ti,ab OR c5ar1:ti,ab) | 2,016 |
| 2 | ( 'antineutrophil cytoplasmic antibody associated vasculitis'/exp OR 'granulomatosis with polyangiitis'/exp OR 'microscopic polyangiitis'/exp OR ('ANCA' NEAR/3 vasculitis):ti,ab OR (granulomatosis NEAR/3 polyangiitis):ti,ab OR wegener*:ti,ab OR (microscopic NEAR/3 polyangiitis):ti,ab OR ('pauci-immune' NEAR/3 glomerulonephritis):ti,ab OR ('rapidly progressive' NEAR/3 glomerulonephritis):ti,ab OR ('renal-limited' NEAR/3 vasculitis):ti,ab) | 39,845 |
| 3 | #1 AND #2 | 656 |
| 4 | Filter: Human | 640 |
| 5 | Date Filter 10/01/2021- 2/28/2026 | 525 |

### **Table S6. PubMed search strategy**

| **Line** | **Search string** | **Hits** |
| --- | --- | --- |
| 1 | (avacopan[tiab] OR tavneos[tiab] OR ccx168[tiab] OR "c5a receptor antagonist"[tiab] OR "c5ar antagonist"[tiab] OR c5ar1[tiab] OR "complement 5a receptor"[tiab] OR "Receptor, Anaphylatoxin C5a"[Mesh]) | 2,008 |
| 2 | ("ANCA-associated vasculitis"[tiab] OR "antineutrophil cytoplasmic antibody-associated vasculitis"[tiab] OR "granulomatosis with polyangiitis"[tiab] OR wegener*[tiab] OR "microscopic polyangiitis"[tiab] OR "pauci-immune glomerulonephritis"[tiab] OR "rapidly progressive glomerulonephritis"[tiab] OR "renal-limited vasculitis"[tiab] OR "Anti-Neutrophil Cytoplasmic Antibody-Associated Vasculitis"[Mesh] OR "Granulomatosis with Polyangiitis"[Mesh] OR "Microscopic Polyangiitis"[Mesh] ) | 20,232 |
| 3 | #1 AND #2 | 238 |
| 4 | Human | 162 |
| 5 | 10/01/2021- 2/28/2026 | 123 |

### **Table S7. Cochrane CENTRAL search strategy**

| **Line** | **Search string** | **Hits** |
| --- | --- | --- |
| 1 | avacopan or tavneos or CCX118 OR CCX-118 or CCX 118 | 92 |
| 2 | 10/01/2021- 2/28/2026 | 65 |
